# The Shift of Percent Excess Mortality during the COVID-19 pandemic (2020 – 2022) in Singapore, South Korea, Australia, New Zealand and Hong Kong SAR

**DOI:** 10.1101/2022.08.31.22279422

**Authors:** Xiaohan Cao, Yunlong Zi, Yuyan Zhu

**Author notes:** **Correspondence:** Yunlong Zi, Yuyan Zhu.

## Abstract

**Introduction:** With the economic recession and pandemic fatigue, milder viral variants and higher vaccine coverage along the time lay the basis for lifting anti-COVID policies to restore COVID-19 normalcy. However, when and how to adjust the anti-COVID policies remain under debate in many countries.

**Methods:** In this study, four countries (Singapore, South Korea, Australia, and New Zealand) and one region (Hong Kong SAR), that have shifted from the zero-COVID (ZC) policy to or close to the living-with-COVID (LWC) during or after the Omicron outbreak, were selected as research objects. All-cause mortality data were collected for these objects from 2009-2019. The expected mortality was estimated by a simple linear regression method. Excess mortality over time was calculated as the difference between the expected mortality and the observed mortality. Finally, percent excess mortality (PEM) was calculated as the excess mortality divided by the expected mortality.

**Results:** In the examined four countries, PEM fluctuated around 0% and was lower than 10% most of the time under the ZC policy before 2022. After shifting to the LWC policy, all the examined countries increased the PEM. Briefly, countries with high population density (Singapore and South Korea) experienced an average PEM of 20-40% during the first half of 2022, and followed by a lower average PEM of 15-18% during the second half of 2022. For countries with low population density under the LWC policy, Australia experienced an average PEM of 39.85% during the first half of 2022, while New Zealand was the only country in our analysis that achieved no more than 10% in average PEM all the time. On the contrary, Hong Kong SAR under their ZC policy attained an average PEM of 71.14% during the first half of 2022, while its average PEM decreased to 9.19% in the second half of 2022 with LWC-like policy.

**Conclusion:** PEM under different policies within each country/region overtime demonstrated that the mortality burden caused by COVID-19 had been reduced overtime. Moreover, anti-COVID policies are suggested to control the excess mortality to achieve as low as 10% in PEM.

**Contribution to the field:**

- This study compared excess mortality within the same country/region, instead of among countries, thus, PEM during the outbreaks of different SARS-cov-2 variants overtime could reflect the effectiveness of regional specific anti-pandemic policies in protecting the lives of citizens locally.
- Our analysis demonstrated that Singapore, South Korea and Australia might implement the LWC policy without sufficient preparation, which resulted in a very high mortality burden during the first half of 2022.
- The reduced PEM in late 2022 in the examined countries/regions suggested that the mortality burden caused by COVID-19 was reduced overtime, laying a great foundation to call for a further relief of LWC policy in the world in the near future.
- This study delineated a threshold of percent excess mortality, which is 10%, as a criterion to assess the effectiveness of anti-COVID policies.

## Introduction

On November 24, 2021, a new variant of SARS-CoV-2 (B.1.1.529) was reported to the World Health Organization (WHO) by South Africa which was later named Omicron (1). Since the emergence of the Omicron variant, the world has entered a post-COVID-19 era. Compared with other variants, Delta in particular, Omicron is characterized by its relatively low pathogenicity and high transmissibility (2). The low pathogenicity considerably reduces the risks of hospitalization and fatality; however, the high transmissibility significantly increases the number of confirmed cases, which in turn, may overwhelm hospitals and cause high mortality in the end. Taken together, whether the mortality burden caused by the Omicron surge is tolerable to the society is unclear. Subsequently, when and how to adjust the anti-COVID policies has been under debate in the world.

Besides the evolution of the SARS-Cov-2 variants, with the accelerated vaccination coverage and the emergence of effective antiviral drugs (3), the case fatality rate of SARS-CoV-2 virus decreased from 80 times higher than that of influenza in April 2020 to less than 2 times higher than that of influenza in early 2022 (4). Accordingly, some countries, such as Singapore and New Zealand, transitioned step by step from a zero-COVID (ZC) policy to a living-with-COVID (LWC) policy prior to or during the Omicron outbreak (5). By contrast, some other countries, such as China,(4) continue to stick to the dynamic ZC policy, with the considerations of the limited medication resources, the high transmissibility of Omicron and its tendency to escape from vaccine-induced immunity (6). The ZC policy aims at zero uncontrolled transmission of COVID-19 viruses in a specific geographic region (5) by means of control measures such as COVID mass testing, case quarantine, contact tracing, and border closure (7) to varying degrees depending on their epidemiological situations. Existing evidence showed that the ZC policy could effectively prevent the spread of the virus and significantly reduce the fatality rate by up to 96% (8). In addition, China kept positive economic growth in 2020 and 2021 under the ZC policy, which was not easy considering the worldwide economic hardship (9). However, the continuation of the ZC policy has its own challenges. Taking China as an example, its current “dynamic ZC” policy is encountering enormous pressure and high costs of disease prevention, especially during the epidemic outbreak in Shenzhen, Jilin Province, and Shanghai in the first half of 2022 (10, 11). In addition, more stringent prevention and control measures during the pandemic affect the quality of life, which is owing to the decreased social connections caused by mandated lockdowns and socially restrictive physical distancing (12). However, when and how to implement the LWC policy appropriately need to consider the balance between the public health and economics.

Mortality rate is considered as an objective indicator to assess the burden of the disease on society and is also the basis for decision-making in the public health (13). The quantification of COVID-associated deaths varies among countries/regions due to the differences in the definition of “COVID-associated deaths”, such as the calculation of the number of cases that “die from COVID-19” and “die with COVID-19” (14). Besides COVID-associated deaths, there might also be non-negligible deaths due to insufficient medical resources during the pandemic, which are not included in the statistics of COVID-19 deaths (14). Thus, COVID-associated deaths alone underestimate the impact of the pandemic. By contrast, all-cause mortality is more robust and objective. To better evaluate the magnitude of COVID-19 and its effects on society, scientists proposed the use of “excess mortality,” which is defined as the net difference between observed mortality and expected mortality (15). The recent mainstream studies (14-16) focused on comparing excess mortality/percent excess mortality (PEM) and COVID-associated deaths/death rate. These comparisons mainly reflected the differences in the measurements of deaths across countries/regions. However, the impact of different virus variants or different anti-pandemic policies on society within the same country/region has not been evaluated. Therefore, there is a lack of evidence to evaluate the effectiveness of the LWC policy in saving lives during the Omicron era overtime.

Herein excess mortality was employed to evaluate the effectiveness of different policies in protecting the lives of citizens within the same country/region during the pandemic. Based on excess mortality, this study adopted the concept of PEM for assessing the mortality burden attributed to different variants of the virus and different public health policies. PEM is the percentage of excess mortality divided by the threshold (15), which is the expected mortality in this study. Since the SARS-COV-2 variant and the vaccination rate were the dominant factors in adopting the LWC policy from the ZC policy in many areas, we selected four countries (Singapore, South Korea, Australia, New Zealand) and one region (Hong Kong) as the representative research objects. The four countries implemented the LWC policy during the Delta/Omicron era, while Hong Kong experienced Omicron outbreaks under specialized ZC policies. The influence of natural fluctuation in expected mortality each year was considered via simple linear regression. This analysis focused on the changes in PEM within the same country/region over the entire pandemic period. Our results delineated a threshold of PEM as a criterion to assess the effectiveness of anti-COVID policies. Furthermore, our study revealed the significant reduction in PEM overtime in the examined countries/regions under LWC policy and suggested that the mortality burden caused by COVID-19 was reduced overtime, laying a great foundation to call for a further relief of LWC policy in the world in the near future.

## Materials and Methods

### 1. Data Collection

Data on all-cause mortality were obtained for four countries (i.e., Singapore, South Korea, Australia, and New Zealand) and one region (i.e., Hong Kong) from governmental sources, including either weekly or monthly mortality data during the pandemic from January 2020 to September 2022 for Singapore, September 2022 (week 39) for South Korea, July 2022 (week 30) for Australia, October 2022 (week 43) for New Zealand and September 2022 for Hong Kong. Details were listed in Table 1. In addition, data on confirmed cases and COVID-associated deaths of four countries and one region were extracted from Google’s COVID map (originally from Johns Hopkins University) (17).

**Table 1.** Source of all-cause mortality data by countries/regions and the time period selection for calculating expected mortality/percent excess mortality.

|  | Source | Time Unit | Time Period Used to Estimate Expected Mortality | Time Period Used to Calculate Percent Excess Mortality | URL |
| --- | --- | --- | --- | --- | --- |
| Singapore | Singapore Deaths By Ethnic Group And Sex, Monthly (20) | Month | January 2009 – December 2019 | January 2020 – September 2022 | https://tablebuilder.singstat.gov.sg/table/TS/M810121 |
| South Korea (i.e. Republic of Korea) | South Korea Human Mortality Database Short-term Mortality Fluctuations (21) | Week | Week 1, 2010 – Week 52, 2019 | Week 1, 2020 – Week 39, 2022 | https://mpidr.shinyapps.io/stmortality/ |
| Australia | Australia Human Mortality Database Short-term Mortality Fluctuations (21) | Week | Week 1, 2015 – Week 52, 2019 | Week 1, 2020 – Week 30, 2022 | https://mpidr.shinyapps.io/stmortality/ |
| New Zealand | New Zealand Human Mortality Database Short-term Mortality Fluctuations (21) | Week | Week 1, 2011 – Week 52, 2019 | Week 1, 2020 – Week 43, 2022 | https://mpidr.shinyapps.io/stmortality/ |
| Hong Kong | Hong Kong Monthly Digest of Statistics (22) | Month | January 2009 – December 2019 | January 2020 – September 2022 | https://www.censtatd.gov.hk/en/EIndexbySubject.html?pcode=B1010002&scode=460 |

### 2. Calculation of expected mortality

Expected mortality is defined as deaths that occurred in a period assuming there is no pandemic and is estimated based on the past trends of all-cause mortality. This study adopted the simple linear regression method to estimate expected mortality. Data on all-cause mortality were obtained for four countries (i.e. Singapore, South Korea, Australia, and New Zealand) and one region (i.e. Hong Kong SAR) from 2009 to 2019. Firstly, we examined different periods of the death data ranging from 2009 to 2019 and employed simple linear regression analysis. Only the data period with R^2^ larger than 0.85 in the linear regression analysis was selected for further calculation (Table 1). Next, within the selected period, linear regression analysis was performed on the data of the corresponding week/month to calculate the expected mortality during the pandemic. For example, the death data of Januarys from 2009 to 2019 in Singapore were analyzed with linear regression to calculate the expected mortality in January 2021 and 2022.

### 3. Calculation of excess mortality and PEM

Based on expected mortality, excess mortality was calculated using the equation below:

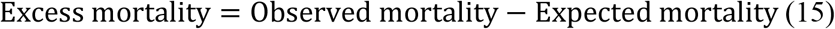

Accordingly, PEM was calculated using the equation below:

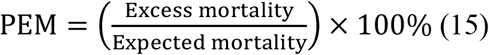

### 4. Calculation of percent COVID-excess mortality (PCEM)

PCEM is defined as the percentage of COVID-associated deaths divided by expected mortality. Data on COVID-associated deaths were obtained from Google’s COVID map (originally from Johns Hopkins University) (17). PCEM was calculated using the equation below:

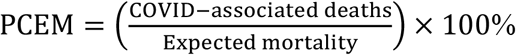

By comparing PCEM with PEM, we can see the contribution of COVID-associated deaths to the overall excess mortality during the pandemic period.

## Results

- **Singapore:** <u>PEM in Singapore under the ZC policy fluctuated around 0% and did not exceed 10%. After shifting to the LWC policy, PEM reached over 10% with an average of 24.23% in response to the Delta variant in late 2021. Then Singapore encountered the Omicron outbreaks. The average PEM was 23.98% in early 2022 and 18.53% in late 2022.</u>

Singapore had a total population of 5.686 million as of 2020 (18) and a population density of 8019 people per square kilometer (19). From January 2020 to July 2021, Singapore effectively implemented the ZC policy, and the total number of deaths from COVID was only 37 (Figure 1A). In August 2021, Singapore announced the implementation of the LWC policy. Since then, it experienced four rounds of COVID-19 outbreaks. The first one was the Delta epidemic from September to December 2021, with a daily increase of more than 3,000 confirmed cases and 10-15 COVID-associated deaths daily at the peak (Figure 1A); the second COVID outbreak was the Omicron epidemic starting from the end of January 2022 to April 2022 with a daily increase of 17,000-19,000 confirmed cases but only about 10 daily COVID-associated deaths at the peak (Figure 1A). Omicron and it evolving variants raised the third (July - September 2022) and fourth outbreak (starting from the end of September 2022) with a daily increase of 5000-12,000 confirmed cases and less than 10 daily COVID-associated deaths (Figure 1A). These above low COVID-associated death data posed Singapore as the world model for handling the COVID pandemic. However, the definition of COVID-associated deaths varies from country to country. When PEM was used to evaluate the total mortality burden under the LWC policy, the analysis, as shown below, suggested a different conclusion.

**Figure 1.**
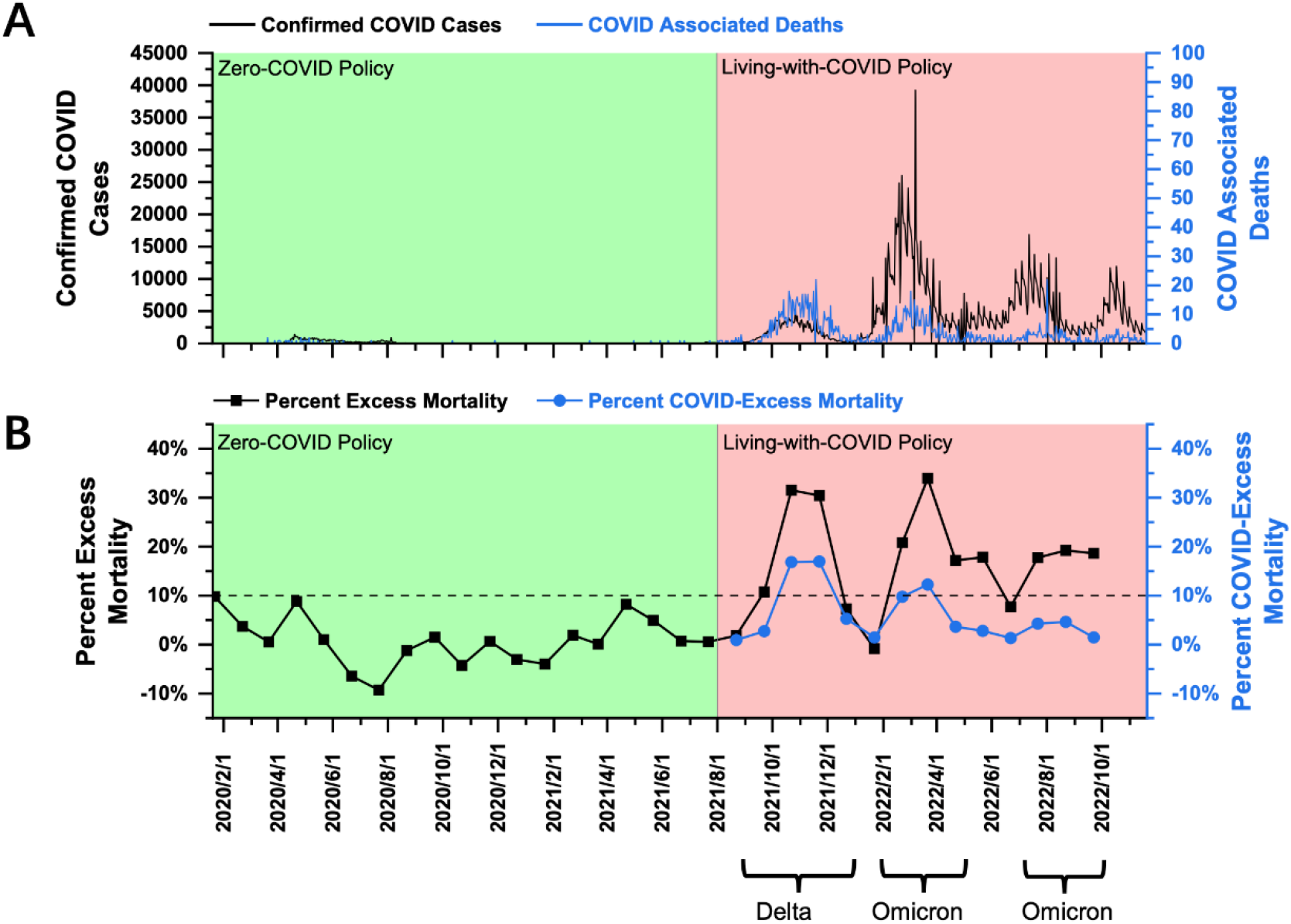
COVID-19 pandemic and mortality statistics in Singapore. (A) SARS-CoV-2 confirmed cases (black) and COVID-associated deaths (blue) from January 22, 2020, to Novemver 20, 2022 (17). (B) Percent excess mortality (monthly) from January 2020 to September 2022 and percent COVID-excess Mortality (monthly) during the living-with-COVID policy period from August 2021 to September 2022. The dotted line is the 10% percent excess mortality/percent COVID-excess mortality line.

Using the monthly mortality data published by the Singapore government (20), a PEM curve from January 2020 to September 2022 was obtained and shown in Figure 1B. PEM in Singapore under the ZC policy before August 2021 fluctuated around 0% and did not exceed 10%. After shifting to the LWC policy in August 2021, Singapore encountered the Delta outbreak. Peak PEM was as high as 31.53% (October 2021), and the average during the Delta outbreak (September - December 2021) was 24.23% (Figure 1B). The Omicron outbreak began in late January 2022. PEM peaked at 33.94% in March 2022, with an average value of 23.98% (February - April 2022) (Figure 1B). The second Omicron outbreak began in mid-June and lasted until September with a peak and an average PEM of 19.23% (August 2022) and 18.53%, respectively (July – September 2022) (Figure 1B). Data on all-cause mortality after October 1, 2022 have not been released by the Singapore government; thus analysis of PEM during the third Omicron Outbreak is out of scope in this paper. The current data showed that PEM increased greatly after the policy transition during both Delta and Omicron outbreaks. In addition, under LWC policy, PEM remained high for Delta and the first Omicron outbreak, while PEM dropped significantly as it progressed into the second half of 2022. Furthermore, PCEM curve was significantly lower than PEM curve (Figure 1B), suggesting a good number of deaths were caused by COVID indirectly under the LWC policy. It may be attributed to the overwhelmed medical resources or the under-quantification of COVID-associated deaths. Collectively, the LWC policy in Singapore in the first half of 2022 failed to control the mortality burden well. However, as the COVID variant evolved and became less virulent, as well as an increased rate of vaccine inoculation and potential development of herd immunity, PEM decreased significantly overtime but remained above 10%.

- **South Korea:** <u>PEM in South Korea under the ZC policy fluctuated around 0%, and most of the time, it did not exceed 10%. After shifting to the LWC policy, PEM exceeded 10% and averaged at 12.83% in response to the Delta variant in late 2021. Then, South Korea faced the Omicron outbreaks with an average PEM of 43</u>.<u>59% in early 2022 and 14.91% in late of 2022.</u>

South Korea had a population of 51.836 million as of 2020 (18) and a population density of 532 people per square kilometer (19). From January 2020 to November 2021, South Korea implemented the ZC policy, and the total number of COVID-19 confirmed cases and the total number of COVID-associated deaths were 368,000 and 2,874, respectively (Figure 2A). On November 1, 2021, South Korea declared to live with COVID. Since then, South Korea experienced a wave of Delta outbreak from November to December 2021, with more than 7,000 daily confirmed cases and 70-80 COVID-associated deaths daily at the peak (Figure 2A). Later from February to May 2022, South Korea faced a wave of Omicron outbreak. The daily confirmed cases were about 400,000, and the daily COVID-associated deaths were 350-400 at the peak (Figure 2A). Moving forward to the second half of 2022, South Korea encountered another Omicron outbreak (July - September 2022) with a daily increase of 5000-15,000 confirmed cases and around 50 daily COVID-associated deaths (Figure 2A).

**Figure 2.**
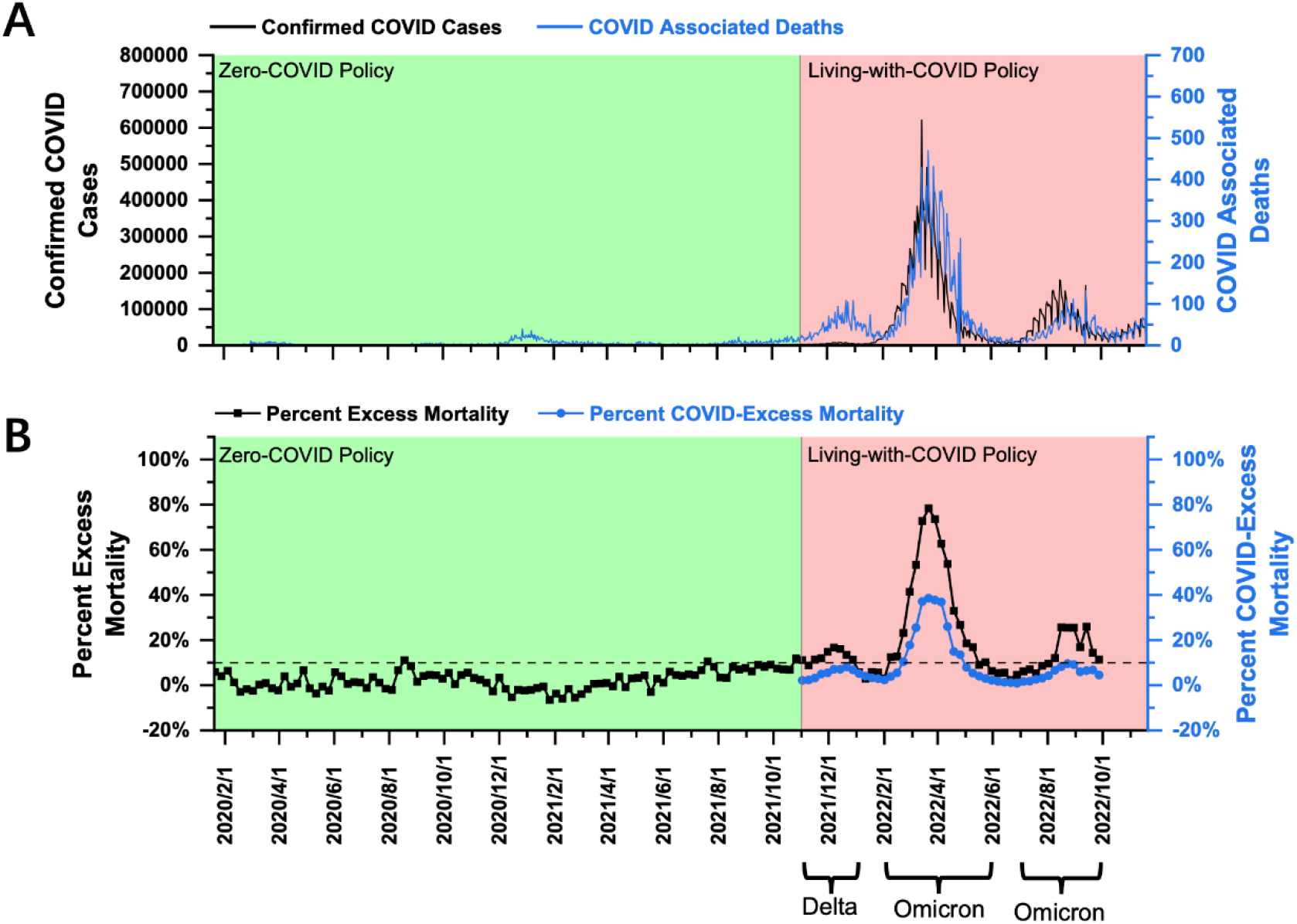
COVID-19 pandemic and mortality statistics in South Korea. (A) SARS-CoV-2 confirmed cases (black) and COVID-associated deaths (blue) from January 22, 2020, to November 20, 2022 (17). (B) Percent excess mortality (weekly) from week 3, 2020 to week 39, 2022 and percent COVID-excess mortality (weekly) during the living-with-COVID policy period from week 44, 2021 to week 39, 2022. The dotted line is the 10% percent excess mortality/percent COVID-excess mortality line.

Using the weekly mortality data in South Korea provided by the Human Mortality Database (which has been collated with the data published by the Korean government) (21), a PEM curve from January 2020 to September 2022 (week 3, 2020 - week 39, 2022) was obtained and shown in Figure 2B. PEM in South Korea under the ZC policy from January 2020 to November 1, 2021 (week 3, 2020 - week 44, 2021) fluctuated around 0%, and most of the time, it did not exceed 10% (Figure 2B). After transitioning to the LWC policy on November 1, 2021, South Korea encountered the Delta outbreak. PEM peaked at 16.70% in December 2021 (week 49, 2021), and the average was about 12.83% in November and December 2021 (weeks 44-52, 2021) (Figure 2B). Statistically, South Korea performed better than Singapore in Delta prevention under the LWC policy. However, considering the high population density in Singapore, the policies in the two countries cannot be compared by statistics only.

PEM in South Korea fell below 0% in January 2022(Figure 2B. It could be partially due to the decline of COVID-associated death with the temporary pandemic recession and possibly due to the limitation in estimating expected mortality for January 2022. Notably, South Korea had a significantly large number of deaths in January 2018, which exceeded 7,000 per week, whereas the number of deaths was less than 6,000 per week in January in years prior to 2018 (21). Thus, a large number of deaths in January 2018 shifted the fitting curve upwards and increased the estimation of expected mortality. Later from February 2018 until the beginning of the pandemic, South Korea’s death toll remained to be similar to that in the previous years. The reason for the sudden increase in the number of deaths in January 2018 remains unknown. If the average number of weekly deaths in January 2019-2021 was used to calculate expected mortality, PEM in January 2022 was still below 10%. Thus, the mortality burden in January 2022 is comparable to that under ZC policy.

The Omicron outbreak occurred from February to May 2022. PEM peaked at about 78.33% in February 2022 (week 12, 2022), and the average was 43.59% from February to May 2022 (weeks 7-19, 2022) (Figure 2B). In the second half of 2022, South Korea experienced another round of Omicron outbreak from July to September 2022 (weeks 27-39, 2022) with a peak and average PEM of 25.93% (week 37, 2022) and 14.91% (Figure 2B). Analysis of PEM in months beyond October 1, 2022 was unavailable until the government’s further release of data on all-cause mortality. The current data showed that PEM increased greatly after the policy transition during both Delta and Omicron outbreaks. In addition, under the LWC policy, the mortality burden decreased significantly in response to two Omicron outbreaks that occurred in the first and second half of 2022. Furthermore, PECM curve was significantly lower than PEM curve during the Omicron outbreak (Figure 2B), suggesting that a good number of deaths were caused by COVID indirectly under the LWC policy when encountering the Omicron variant. It might be attributed to the overwhelmed medical resources or the under quantification of COVID-associated deaths. Collectively, the LWC policy in South Korea in early 2022 failed to control the mortality burden well. However, later in the second half of 2022, the average PEM, although it remained above 10%, dropped drastically. Decreasing mortality burden might be due to less virulent variants and the potential development of herd immunity.

- **Australia:** <u>PEM in Australia under the ZC policy fluctuated around 0%, and most of the time, it did not exceed 10%. After shifting to the LWC policy, Australia encountered two subsequent Omicron outbreaks. PEM reached an average of 39.85% in early 2022 and decreased to 35.68% in late 2022.</u>

Australia had a population of 25.693 million as of 2020 (18) and a population density of 3 people per square kilometer (19). From January 2020 to October 11, 2021, Australia effectively implemented the ZC policy. The total confirmed cases of COVID-19 were 131,000, and the total COVID-associated deaths were 1,461 (Figure 3A). On October 11, 2021, Australia announced the beginning of the LWC policy, which coincided with the Delta outbreak, leading to a daily increase of more than 2,000 confirmed cases and daily COVID-associated death of 10-15 (Figure 3A). Since then, Australia has experienced two waves of Omicron outbreaks. The first wave lasted from the end of December 2021 to March 2022, with more than 100,000 confirmed cases per day (Figure 3A) and more than 80 COVID-associated deaths per day. The second wave started at the end of February 2022, while the first wave had not completely subsided until August 2022. During the second wave of Omicron, there was a daily increase of more than 50,000 confirmed cases and a large fluctuation of daily COVID-associated deaths of about 20-50 (Figure 3A).

**Figure 3.**
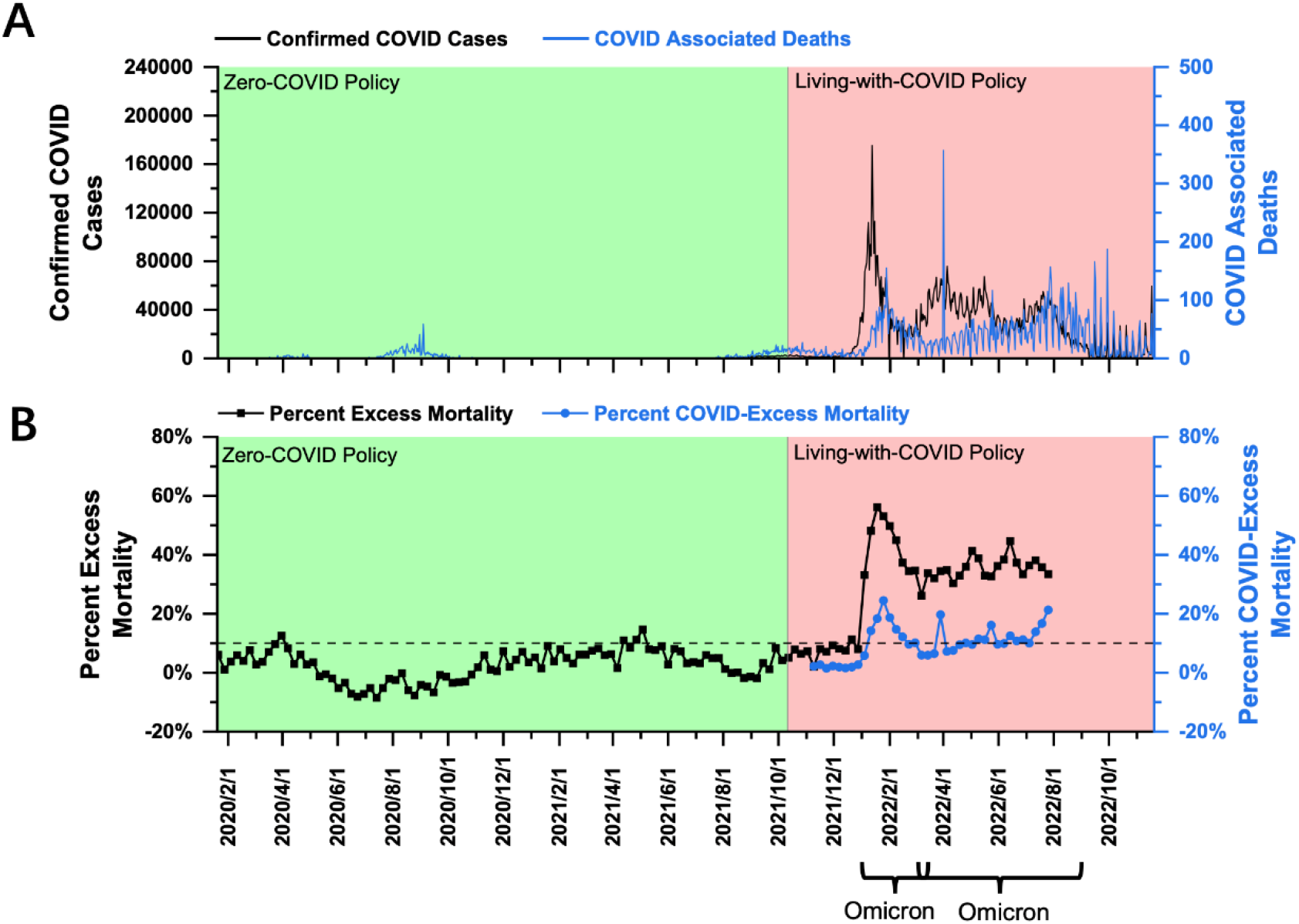
COVID-19 pandemic and mortality statistics in Australia. (A) SARS-CoV-2 confirmed cases (black) and COVID-associated deaths (blue) from January 22, 2020, to November 20, 2022 (17). (B) Percent excess mortality (weekly) from week 3, 2020 to week 30, 2022 and percent COVID-excess mortality (weekly) during the living-with-COVID policy period from week 40, 2021 to week 30, 2022. The dotted line is the 10% percent excess mortality/percent COVID-excess mortality line.

Using the Australian weekly death data provided by the Human Mortality Database (which has been collated with the data published by the Australian government) (21), a PEM curve from January 2020 to July 2022 (week 3, 2020 – week 30, 2022) was obtained and shown in Figure 3B. Australia only published data on all-cause mortality for 2015-2019, and the annual death data fluctuated significantly, so it was impossible to perform any effective linear fitting. Therefore, in this study, the average number of yearly deaths for 2016-2019 was used to estimate expected mortality. As shown in Figure 3B, PEM in Australia under the ZC policy before October 11, 2021 (week 40, 2021) fluctuated around 0% and was below 10% most of the time. After transitioning to the LWC policy on October 11, 2021, Australia experienced a Omicron outbreak beginning in late December 2021, and PEM rose to nearly 10% in the last two weeks of 2021. As Australia continued its LWC policy, the impact of Omicron was significantly enhanced, with a peak PEM at about 56.12% in January 2022 (week 3, 2022) and an average of 39.85% from January and March 2022 (weeks 1-13, 2022). The second wave of the Omicron outbreak (March - August 2022) achieved a peak PEM of 44.65% (Week 24, 2022) and an average PEM of 35.68% (Weeks 11-30, 2022) (Figure 3B). The second Omicron outbreak lasted until the end of August 2022. However, all-cause death data beyond August 1, 2022, was not released by the government, so PEM analysis in this study covered up to July 2022. The current data showed that PEM increased significantly after the policy transition during two waves of the Omicron outbreak. In addition, under the LWC policy, average PEM decreased in response to two Omicron outbreaks. Furthermore, PCEM curve was significantly lower than PEM curve (Figure 3B), suggesting that a good number of deaths were caused by COVID indirectly under the LWC policy. It might be attributed to the overwhelmed medical resources or the under quantification of COVID-associated deaths. Collectively, the LWC policy in Australia failed to control the mortality burden well. Average PEM decreased slightly in response to two subsequent Omicron outbreaks that occurred in early and late 2022, which might be due to a decreasing virulence of the virus and the potential development of herb immunity through acquired immunity and vaccine inoculation.

- **New Zealand:** <u>PEM in New Zealand under the ZC policy fluctuated around 0%, and most of the time, it did not exceed 10%. After shifting to the LWC policy, New Zealand encountered Omicron outbreaks. PEM averaged at 9.48% and 7.67% in early and late 2022, respectively.</u>

New Zealand had a population of 5.090 million as of 2020 (18) and a population density of 19 people per square kilometer (19). From January 2020 to November 2021, New Zealand effectively implemented the ZC policy. The total confirmed cases were nearly 120,000, and the total COVID-associated deaths were 44 during this ZC policy period (Figure 4A). On December 3, 2021, New Zealand implemented the LWC policy. Then it encountered two waves of Omicron outbreak. The first began in February 2022 and lasted until the end of April 2022, with more than 20,000 daily confirmed cases (Figure 4A) and 10-20 daily COVID-associated deaths. The second Omicron outbreak (June - August 2022) experienced 5000-10,000 daily confirmed cases and about 10 daily COVID-associated deaths (Figure 4A).

**Figure 4.**
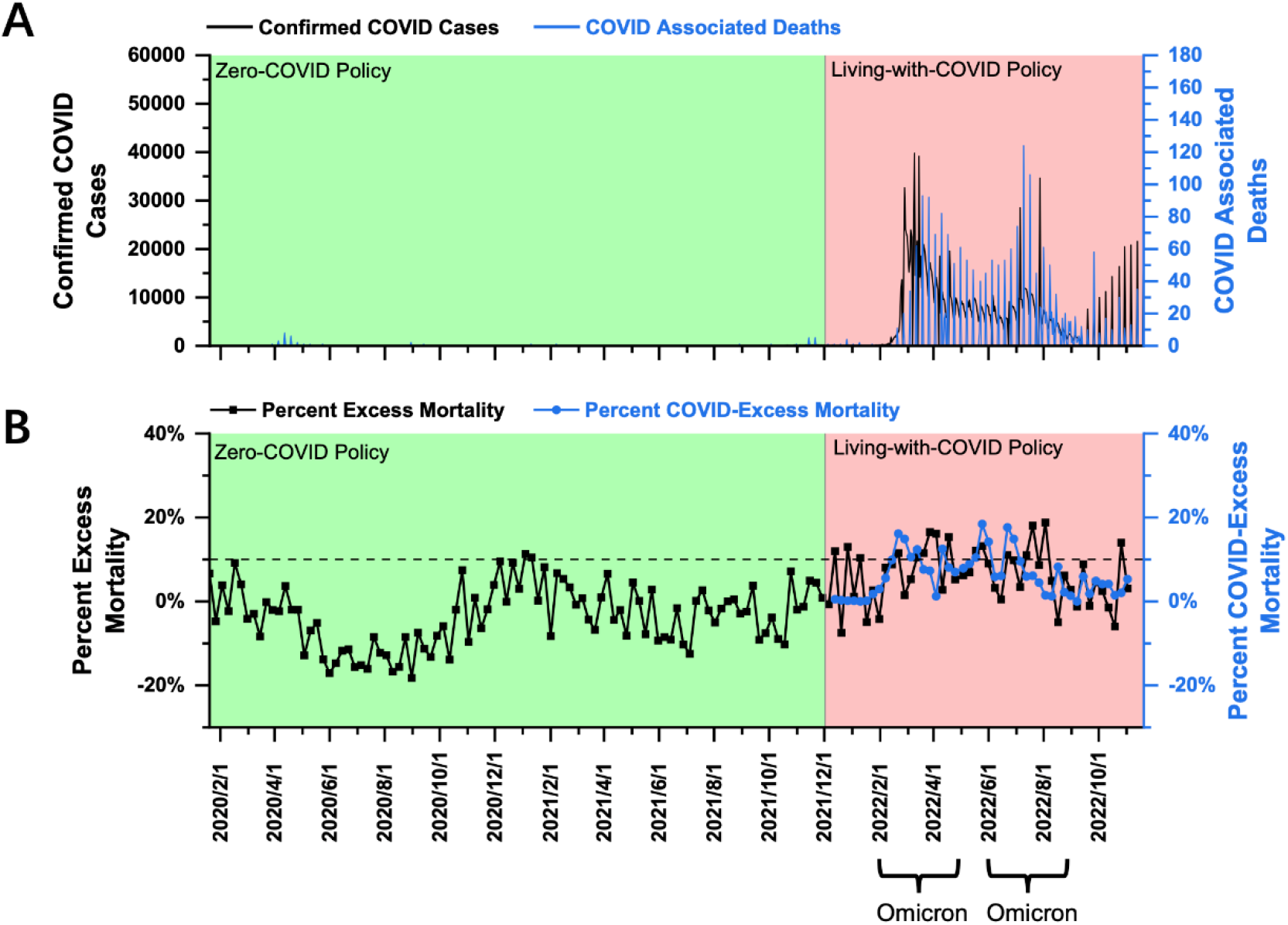
COVID-19 pandemic and mortality statistics in New Zealand. (A) SARS-CoV-2 confirmed cases (black) and COVID-associated deaths (blue) from January 22, 2020, to November 20, 2022 (17). (B) Percent excess mortality (weekly) from week 3, 2020 to week 43, 2022 and percent COVID-excess mortality (weekly) during the living-with-COVID policy period from week 47, 2021 to week 43, 2022. The dotted line is the 10% percent excess mortality/percent COVID-excess mortality line.

Using the weekly death data in New Zealand provided by the Human Mortality Database (which has been collated with the data published by the New Zealand government) (21), a PEM curve from January 2020 to October 2022 (week 3, 2020 - week 43, 2022) was obtained and shown in Figure 4B. PEM under the ZC policy before December 2021 fluctuated around 0% and remained below 10% (Figure 4B). After transitioning to the LWC policy on December 3, 2021, New Zealand experienced a relatively long period of a steady phase from December 2021 (week 47) to late-February 2022 (week 9) until the beginning of the Omicron outbreak. Starting in week 10 in 2022, PEM exceeded 10% and continued to rise to a peak at about 16.53% in week 12. The average PEM during the Omicron outbreak (weeks 8-20, 2022) was 9.48% (Figure 4B). The second Omicron outbreak began in June 2022 and lasted until August 2022, and the peak PEM was 18.77% (Week 30, 2022), and the average PEM was only 7.67% (Weeks 21-33, 2022) (Figure 4B).

In our analysis, New Zealand is the only country that achieved approximately 10% average PEM during the Omicron outbreak under the LWC policy, which might be a result of the ultrahigh vaccination rate, especially among the elderly. There was no significant difference between PCEM and PEM curves (Figure 4B), suggesting that data on COVID-associated deaths well reflected the mortality burden attributed to COVID-19. Collectively, the LWC policy in New Zealand in the examined period acceptably succeeded in controlling the mortality burden.

- **Hong Kong:** <u>PEM under the ZC policy before 2022 fluctuated and remained approximately about 10% most of the time. However, during the outbreak of the Omicron variant, the average PEM was 71.14% in the first half of 2022 and decreased to 9.19% in the second half.</u>

Hong Kong had a population of 7.481 million as of 2020 (18) and a population density of 7,126 people per square kilometer (19). Hong Kong has implemented the ZC policy since January 2020. Before Omicron struck at the beginning of February 2022, Hong Kong underwent a period of a stationary state of the pandemic with total confirmed cases of nearly 15,000 and total COVID-associated deaths of 213 (Figure 5A), proving the effectiveness of the ZC policy. Then, from March to the mid-May 2022, Hong Kong experienced a major wave of Omicron outbreak, with over 1.2 million confirmed cases and more than 9000 COVID-associated deaths total (Figure 5A). Then Hong Kong experienced a second wave in August and September 2022, with 400,000 confirmed cases and more than 600 COVID-associated deaths total (Figure 5A).

**Figure 5.**
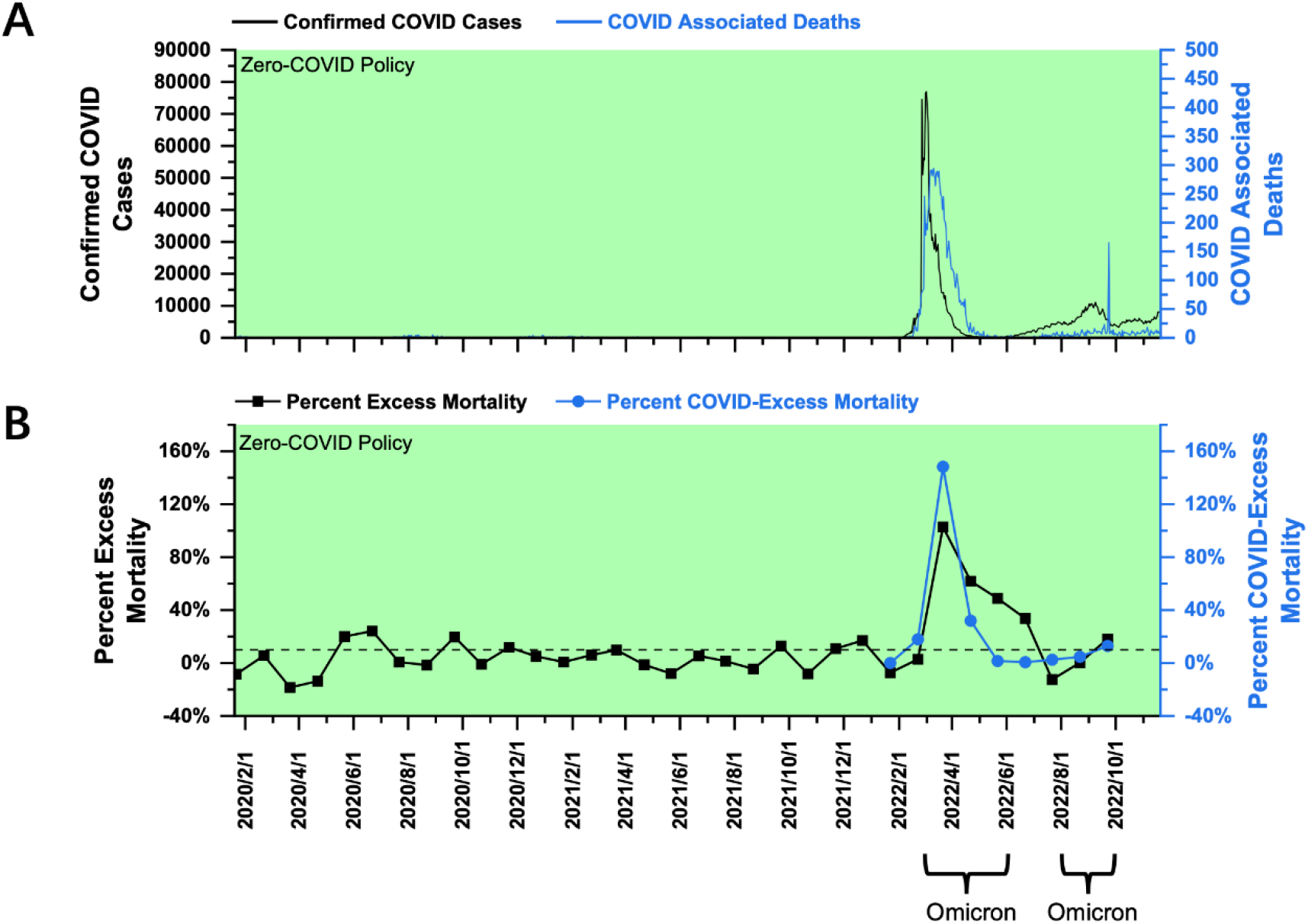
COVID-19 pandemic and mortality statistics in Hong Kong. (A) SARS-CoV-2 confirmed cases (black) and COVID-associated deaths (blue) from January 22, 2020, to November 20, 2022 (17). (B) Percent excess mortality (monthly) in Hong Kong from January 2020 to September 2022 and percent COVID-excess mortality (monthly) from January 2022 to September 2022. The dotted line is the 10% percent excess mortality/percent COVID-excess mortality line.

Using the monthly death data published by the Hong Kong government for analysis (22), a PEM curve from January 2020 to September 2022 was obtained and shown in Figure 5B. PEM under the ZC policy before February 2022 fluctuated and remained approximately about 10% most of the time. The periods with about 20% PEM mainly corresponded to several waves of SARS-CoV-2 in the first two years of the pandemic. Considering the high population density in Hong Kong, the containment measures against COVID-19 executed by the government before February 2022 were acceptable. However, a major wave of the Omicron variant breached the long streak of the stationary phase with a peak PEM of 102.77% in March 2022 and an average PEM of 71.14% (March-May 2022) (Figure 5B). Then, in August and September, Hong Kong experienced another wave of Omicron outbreak with a peak and an average PEM of 18.17% (September 2022) and 9.19% (August -September 2022) (Figure 5B). Data on all-cause mortality from October 1, 2022, onwards were still unavailable from the government website. Even under the ZC policy, Hong Kong failed to control the spread of the Omicron variant in early 2022. The Omicron surge could be attributed to its high population density, the low vaccination rate among the elderly (23), the highly contagious variant, and increased social mixing during the lunar new year. The outbreak in Hong Kong aroused endless debates on the ZC policy and its effectiveness in preventing a variant as contagious as Omicron. The current data showed that PEM increased greatly in response to the Omicron outbreak in early 2022. However, PEM decreased significantly in late 2022 when encountering another wave of Omicron outbreak. Furthermore, PCEM curve was considerably higher than PEM curve in February and March 2022 (Figure 5B), suggesting that a good number of reported COVID-associated deaths might be overreported. It might be because Hong Kong counted patients who “died with COVID” into the pool of “COVID-associated deaths.” Then, PEM curve became higher than PCEM curve, indicating an under quantification of COVID-associated deaths. Collectively, the specialized ZC policy in Hong Kong was unsuccessful in controlling the mortality burden during the Omicron outbreak in the first half of 2022. Average PEM decreased as progressed into the second half of 2022 due to the reduced pathogenicity of the virus and the potential development of herd immunity.

## Discussion

The goal of this study is to delineate a threshold of PEM as a criterion to assess the effectiveness of different anti-pandemic policies in response to different variants of viruses. During the implementation of the ZC policy, PEM in various countries did not exceed 10% most of the time. Occasionally it fluctuated around 10% for a short time and then quickly declined. After shifting to the LWC policy, PEM increased significantly and exceeded 10% in early 2022. Detailed summary of PEM data for each country/region is listed in Table 2. Thus, when PEM is kept roughly at or below 10%, the mortality burden during the COVID pandemic could be considered acceptable to the public, government officials, and healthcare professionals, etc. Therefore, the PEM threshold of 10% might be set as a criterion to assess the effectiveness of anti-pandemic policies in controlling the mortality burden. Furthermore, the policymakers are suggested to control the PEM within 10% during outbreaks.

**Table 2.**
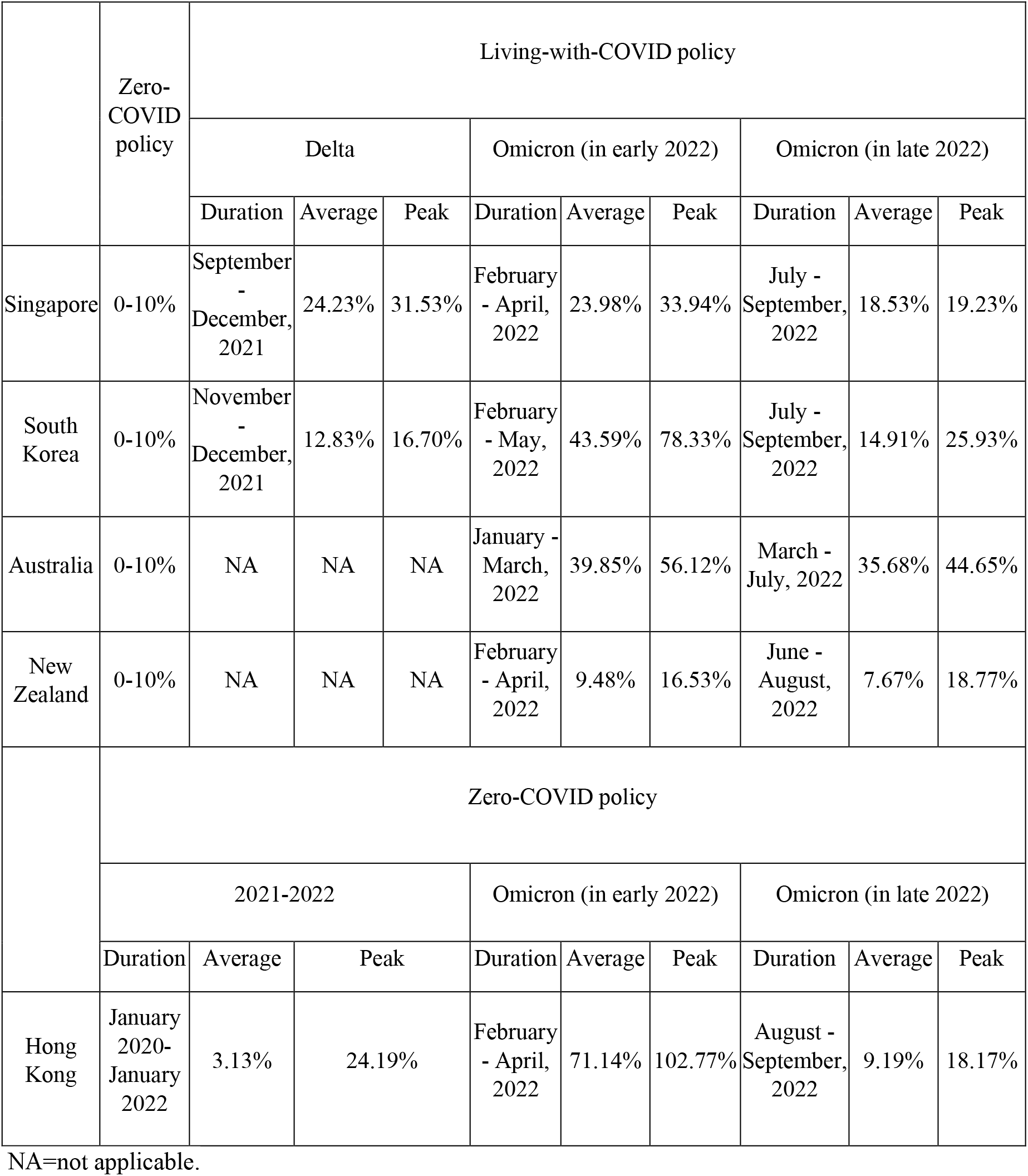
Summary of percent excess mortality stratified by the zero-COVID policy and the living-with-COVID policy, including peak and average percent excess mortality for SARS-CoV-2 Delta and Omicron variants and their corresponding durations for four countries and one region.

The four countries analyzed in this study can be classified into two groups: Singapore and South Korea, which are high in population density, influenced by Confucian culture, and are located in the northern hemisphere, as group A; while Australia and New Zealand, which are low in population density, influenced by Anglo-Saxon culture and are located in the southern hemisphere, as group B. Countries in group A dealt with both Delta and Omicron variants while those in Group B only dealt with the Omicron variant. Peak PEM in Group A was 16-30% and 30-70% in response to the Delta and the Omicron variants, respectively. Regarding countries in Group B and their responses to the Omicron variant, Australia had a PEM of about 10% at the beginning of the outbreak, which later exceeded 10% and reached over 50%. New Zealand maintained a low average PEM of almost 10%, with a peak of 16.69%. Collectively, it is relatively challenging for Group A countries to control the PEM below 10% during the Omicron era. From the mortality burden standpoint, Group A countries/regions may implant the LWC policy without sufficient preparation, which will lead to a high mortality burden in early 2022. While for Group B countries, New Zealand set a good model of exercising LWC policy during omicron outbreaks, which could be learned by other countries/regions with low population density.

PEM during the omicron outbreak was not less than that during Delta outbreaks in Group A countries (Figure 1B, 2B). The difference in PEM in response to the Omicron variant between Group A and New Zealand was mainly due to differences in population density (24). In addition, the higher PEM in Group A countries can be largely attributed to the following factors. First, influenced by the Confucian culture, multigenerational households are more common, which leads to over-crowdedness and increase the risk of COVID-19 transmission and infection (25). Second, the northern hemisphere was experiencing winter during the Omicron outbreak, which was associated with a surge of COVID-19 infections and deaths (26). Such association can be attributed to biological factors, including the susceptibility of COVID viruses to heat and UV-radiation (27, 28), and behavioral factors, such as the tendency to have more gatherings in crowded indoor areas in cold weather.

Hong Kong has been implementing a ZC policy throughout the entire period of the pandemic. The spread of the virus was well controlled in the first two years of the pandemic until the Omicron outbreak. The surge of Omicron in Hong Kong was partially due to low vaccination coverage, especially among the elderly population. By December 23, 2021, 52% of vaccine-eligible individuals received at least one dose, and 49% received at least two doses, of which only 7% received a booster dose for those aged 60 years and above, (23). 96% of COVID-associated deaths during January 6 – March 21, 2022, happened to those elders aged ≥ 60 years, while 70% of this age group were unvaccinated (23). In contrast, New Zealand, as the only country with an average PEM of 10% during the Omicron outbreak under the LWC policy in our analysis, benefited from the ultrahigh vaccination rate, especially among the elderly (96.5% were fully vaccinated for elders above 60 years old and > 90% were boosted for these above 70 years old) (29) (Data were extracted on May 3, 2022). In agreement with multiple studies, a high vaccination rate is associated with low excess mortality and is an essential indicator of adjusting anti-pandemic policy (4, 30). The underlying reasons for the low vaccination in Hong Kong rate remained unclear. Still, it was possibly due to vaccine hesitancy caused by the inefficiency in vaccination promotion and the widespread misunderstandings of the side-effects of COVID vaccines (23, 31). In addition, the Chinese Lunar new year, which coincided with the Omicron outbreak, facilitated the spread of the virus owing to increased gatherings among families and friends. Also, although both were named the “ZC policy,” the precautionary measures implemented by Hong Kong were different and less strict than those in mainland China, which might also fail to control the Omicron surge.

Since the emergence of the Omicron variant, there have been endless debates on public health policies. As most countries transitioned to the LWC policy, countries/regions that insisted on the ZC policy were thrust into the limelight. Numerous parties have criticized government officials’ authoritarian rules imposed on the general public and blamed that the ZC policy lacked basic humanity without considering the specific local conditions, such as population density, population structure, vaccination coverage, availability of healthcare resources, and culture, etc. To ease the evaluation process, we proposed a PEM threshold of 10% as a standard to assess the effectiveness of any anti-pandemic policy from the perspective of whether the mortality burden of the pandemic was tolerable to the society. The analysis of PEM was simple yet efficient and provided a preliminary evaluation of the feasibility of the policy. Although collectively named the LWC policy, the content and stringency highly varied among countries/regions. Thus, it is not a matter of implementing which policy but rather of rules and regulations that can effectively minimize the mortality burden on society during the pandemic. We suggest that anti-pandemic policies be adjusted to achieve a PEM of 10%.

In addition, we found that PEM during Omicron outbreaks in early 2022 was not less than that during Delta outbreaks, suggesting that Omicron should not be the key reason for the policy transition towards LWC. However, PEM decreased significantly from early to late 2022 in all studied countries/regions, suggesting that the mortality burden in response to the Omicron outbreak fell, which might be due to the following factors. First, herd immunity was developed along the pandemic’s progression as more individuals acquired immunity through infection or vaccination (32). Second, the COVID virus variant evolved with reduced pathogenicity, significantly decreasing deaths and severe cases (33). Third, the government was more prompt in adjusting anti-pandemic policies, including enhanced regulation on infectious individuals, to encounter each COVID outbreak. The mortality burden caused by COVID-19 was reduced over time, laying a great foundation to call for further relief of LWC policy in the near future. In the long run, the world may eventually have to coexist with the COVID virus. Still, the preconditions of transitioning to a LWC policy need to be examined, and high vaccination coverage is a crucial requirement. PEM can serve as a reference, but further research to identify more dimensions to assess the impact of the disease is needed for better policy-making and implementation.

There are some limitations in this study. First, PEM is calculated using the all-cause mortality reported by official statistics. Therefore, the practicality and validity of PEM-based analysis depend on the infrastructure and capacity to record and report mortality in each location. In addition, the frequency of data reporting affects the accuracy of data as monthly death reports tend to even out any fluctuations in weekly death reports and result in underestimation. Second, to simplify the evaluation, we did not consider the impact of the economy on health and mortality in the long run. Studies from other angles like economy and life quality are needed to help develop appropriate policies during the pandemic.

## Data Availability

All data produced in the present study are available upon reasonable request to the authors

## Patient and Public Involvement

This work did not have any involvement.

## Abbreviations

PEM: Percent excess mortality
PCEM: Percent COVID-excess mortality
ZC: Zero-COVID
LWC: Living-with COVID

## Author contribution statement

Xiaohan Cao:data collection, literature search, figures, data interpretation, writing; Yunlong ZI: study design, data collection, data interpretation, writing; Yuyan ZHU: study design, literature search, figures, data interpretation, writing.

## Notes

The authors declare no competing financial interest.

## Acknowledgements

We thank Dr. GAO Qi from the Chinese University of Hong Kong and Prof. LU Jian from City University of Hong Kong for the insightful comments and suggestions. Part of data and the manuscript has been released as a pre-print at https://www.medrxiv.org/content/10.1101/2022.08.31.22279422v1 (34).

